# Optimal targeting of interventions uses estimated risk of infectiousness to control a pandemic with minimal collateral damage

**DOI:** 10.1101/2023.10.06.23296661

**Authors:** James Petrie, Joanna Masel

## Abstract

In this paper, we present a simple model that shows how to optimally target interventions based on the estimated risk of infectiousness of individuals. Our model can help policymakers decide when to use different types of interventions during a pandemic, depending on their precision, which is the fraction of positive predictions that are true positives. We show that targeted interventions, even with very low precision, can impose a much smaller overall burden on the population than non-targeted alternatives, such as lockdowns or mass testing. To illustrate this, we use data from the NHS contact tracing system in the UK to construct a risk function based on second degree contact tracing, which is similar to the strategy used by Vietnam in 2020. We find that with moderate precision (greater than 1/1000) and sufficient sensitivity (greater than 1 − 1*/R*_0_), countries can cope with a large number of imported cases without resorting to social distancing measures, while keeping the per-person probabilities of both infection and quarantine very low. We also show that targeted strategies are often orders of magnitude better than default strategies, making them robustly beneficial even under significant uncertainty about most parameters.

## 2 Introduction

Pandemics impose a huge burden, both directly on health, and indirectly via the cost of interventions used to contain them. In future pandemics, effective vaccines and medication may not be immediately be available. In these cases, non-pharmaceutical interventions (NPIs) may be the only disease containment option for months or years, whether implemented by government decree or spontaneously by individuals. Several studies have investigated the use of optimal control to most effectively contain pandemics [1, 2, 3, 4], but they focused on the optimal timecourse of population-wide interventions, where all options are costly. For this reason, Camelo et al. analyzed the approach of targeting social distancing based on age and activity, with the goal of reducing the chance of infection amongst those most likely to experience severe disease [5].

Targeting can focus on those most at risk of infecting others, rather than those most at risk of severe outcomes if infected. Birge et al. modeled the targeted closure of neighborhoods in New York based on local disease prevalence [6]. Abdin et al. considered allocation of tests among compartments (e.g. symptomatic vs. asymptomatic individuals, or regions) on the basis of potential to stem onward transmission [7]. Using the estimated risk of infectiousness can achieve the greatest reduction in transmission with the lowest burden inflicted by control measures on the population. A qualitative version of this logic underlies several standard practices, including testing and quarantine of individuals identified through contact tracing or symptoms. Explicit modeling of this general approach is useful because it reveals when exactly it is helpful to use each type of targeted intervention (based on estimated risk of infectiousness and disease prevalence). Furthermore, it reveals the potential for enormous reductions in the cost of disease control using only moderately informative estimates of infectiousness risk -which could potentially be achieved with existing technology and infrastructure in many countries.

In [8], we evaluated how the risk of infectiousness of a given individual could be used to make optimal quarantine decisions at the margin. In this manuscript, we build a simplified model for an entire population, showing how disease control capabilities depend on the available distribution of estimated infectiousness (for example, as a function of symptoms, test results, contact tracing, geographic base rate or profession). We will show that the per capita cost of a targeted disease control strategy is proportional to the number of infected individuals, rather than proportional to the total population. Since disease prevalence can vary over many orders of magnitude (10^0^ to 10^−9^) in a large population, so can the per capita cost of disease control. Here cost means the total burden of both control measures and disease on the population, in units of quality-adjusted life years [9] or similar. Because quantities vary over such a scale, even very imprecise information (e.g. about disease state, control costs, and population risk) can be used to perform order of magnitude estimates to produce highly actionable recommendations.

We frame the decision of how to make targeted recommendations for social distancing as an optimization problem making use of estimated risk of infectiousness for individuals within the population. The objective function of this optimization problem depends on the cost of social distancing for a single individual as a function of the degree of distancing. We show that when this cost function is either linear or strongly convex, the optimization problem can be simplified and solved efficiently.

For simplicity we assume that individuals are either infectious or not, but the analysis in this manuscript is also valid (and more accurate) if risk of being infectious is replaced with expected transmissions from each individual. A method of classifying infectiousness risk can be summarized based on its precision (True Positives / (True Positives + False Positives)) and sensitivity (True Positives / (True Positives + False Negatives)). Note that correctly detecting all infectious individuals 3 days into a 10 day infectious period would correspond to a sensitivity of 70%. Our assumption removes the complication of timing, in which some days of peak infectiousness are more important than others. In Section 5 we show how sensitivity and precision impact the minimum cost needed to achieve a certain level of disease control.

As an example, consider a country with a population of 100M dealing with 100 daily importations of a pathogen with *R*_0_ = 3 and an infectious period of 5 days. With no controls, the cumulative per-person probability of infection would rise to around 70% ^1^, with most people infected at a time during which medical resources are unavailable. With only broad controls, a persistent 76% reduction^2^ in social contact would be required to reduce the annual chance of infection to 1/1000 during the wait for a vaccine or effective treatment. In comparison, an intervention with 1% precision and 80% sensitivity could achieve the same 1*/*1000 annual chance of infection at an expected cost of 0.4 days in quarantine per person per year, i.e. 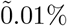 temporally focused reduction in social contact. Either increasing the precision to 10% or decreasing the daily importations to 10 would reduce the cost to 0.04 expected days of quarantine per year.

## 3 Model

### 3.1 Risk Function

Figure 1 shows how we construct a function describing the distribution of estimated relative infectiousness in a population. This normalized risk as a function of quantile function, *f* (*q*), will always be positive, monotonically decreasing, and integrate to 1. It could take a variety of forms depending on the information used, sophistication of risk analysis, as well as specifics of the population and disease. In Section 5 we will investigate how the shape of this function (specifically the concentration of infection risk) influences the cost of disease containment.

**Figure 1.**
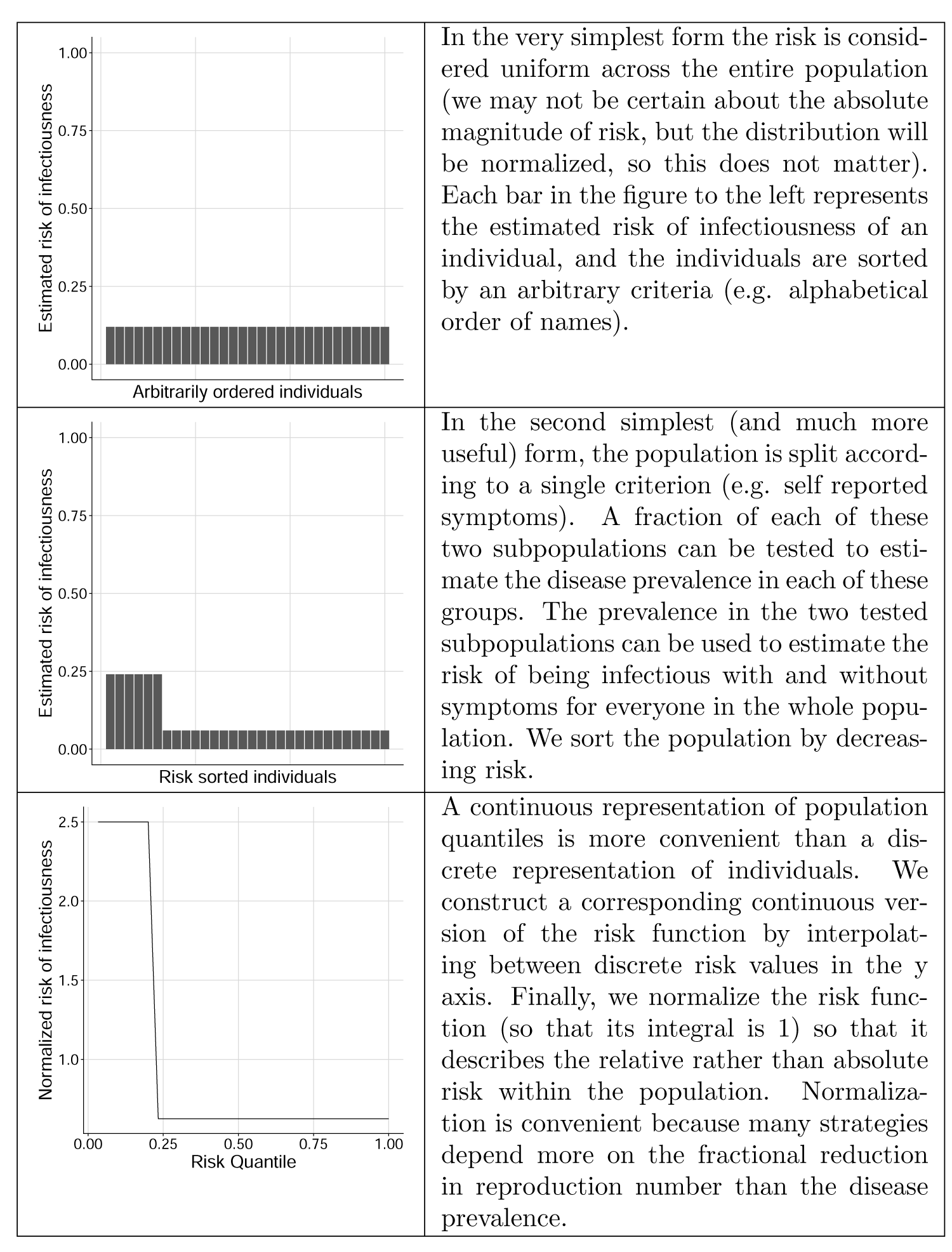
Example steps to construct infection risk function.

### 3.2 Targeted Control of Transmission

In this model, we consider the ability to target interventions towards different portions of the population. For example, at-home isolation is a control policy targeted at particular individuals. The function *D*(*q*) represents the effect of the targeted policy as the fractional reduction of expected transmissions from an infected person who is at risk quantile *q*. Assuming the risk estimator is unbiased, the population-wide fractional reduction in transmissions, *β*, caused by the control policy is:

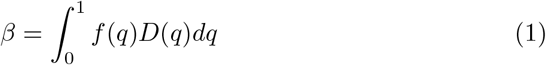

And the modified reproduction number (assuming negligible population immunity) is:

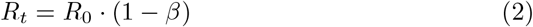

### 3.3 Dynamics

Assuming the generation time, *τ*, is roughly unchanged by applied interventions, the Malthusian growth rate, *r*_*t*_ can be computed as shown in Equation 3 [10].

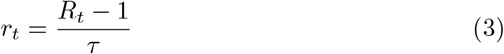

From Equation 2, we have *R*_*t*_ = *R*_0_(1 − *β*) (assuming that the majority of the population remains susceptible^3^). With an import rate of *A* cases per unit time divided by the total population size, the infection dynamics are given by Equation 4.

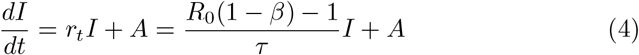

Here *I*(*t*) ∈ [0, 1] is the fraction of the population that is infected (and infectious) at time *t*.

## 4 Minimizing Cost of Disease Control

For a given value of *R*_*t*_, it is clearly beneficial to choose the control strategy that minimizes the burden on the population. Because we’ve chosen to fix *R*_*t*_, health outcomes are constant and so their cost doesn’t need to be considered. As in [8], we assume there is a convex function, *Cost*(*d*), describing the perperson-day cost of reducing transmissions by fraction *d*. The policy choice can be expressed as the following optimization problem over potential transmission reduction functions, *D*(*q*) ∈ [0, 1] → [0, 1] (with *β* = 1 − *R*_*t*_*/R*_0_ as before) :

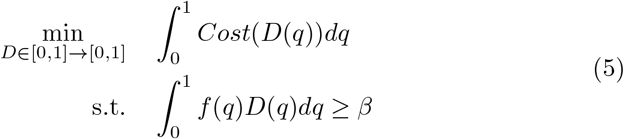

If *Cost*() is an increasing linear function, then an optimal strategy^4^ is given by Equation 6 (Proof in Appendix A). Here 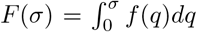 is the integral of the risk function. This solution applies all interventions to the highest risk fraction of the population until the required reduction in transmissions has been achieved.

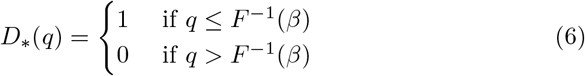

If *Cost*() is a strongly convex function (i.e. if twice the reduction in social contact incurs much more than twice the cost to the individual) then the optimization problem over the space of functions *D*() can be reduced to optimization of a single scalar value, *µ*_*β*_. The proof for this is in Appendix B and the easier form of Equation 5 is:

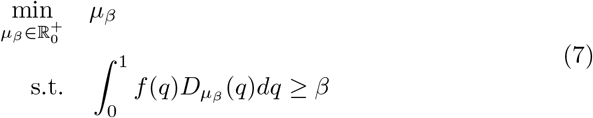

With 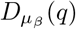 defined as:

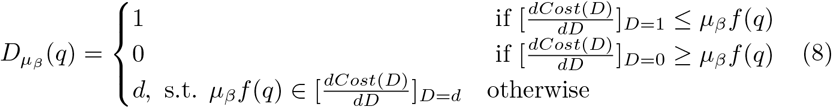

This solution essentially applies maximum control to any portion of the population exceeding a risk threshold, no control to any portion below another risk threshold, and intermediate levels of control depending on risk and the slope of the cost function for the remainder. Both solutions 6 and 7 can be numerically solved very efficiently.

## 5 Dependence on Sensitivity and Precision

Many NPIs and other interventions can be characterized by their precision (True Positives / (True Positives + False Positives)) and sensitivity (True Positives / (True Positives + False Negatives)). As an example, a mass testing strategy that detects half of all infected individuals during half of their infectious window would have a sensitivity of 25%. The precision depends on the number of non-infected contacts multiplied by the number of days that they meet the same criteria (per true positive). Early contact tracing studies for COVID-19 found a secondary attack rate (SAR) of 1% amongst casual contacts [11]. If strategies like testing to reduce quarantine time are not used, then the precision would be slightly lower than this SAR (because all contacts would be recommended to quarantine for longer than the infectious period). Several other strategies could similarly be characterized using precision and sensitivity: for example, quarantining residential areas based on wastewater surveillance might have (at worst) a precision equal to the inverse of the wastewater catchment size and a sensitivity depending on the reliability (and timing) of detection.

Given the sensitivity and precision, a simplified version of the risk function can be constructed as a piece-wise constant function with two categories, as in Equation 9.

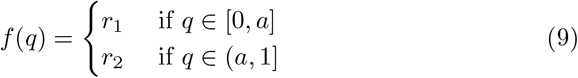

The parameters *a, r*_1_, and *r*_2_ are initially unknown, but can be solved for using the disease prevalence *I*, sensitivity *γ*, and precision *ρ*. Equation 10 describes the proportion *a* classed as “positive”, a fraction proportional to *r*_1_ of which are True Positives. The normalization of the risk function to 1 conveniently makes the product *a* · *r*_1_ equal to the sensitivity *γ*. Equation 11 specifies that the ratio of true positives (*Iγ*) to true and false positives (*a*) equals the precision (*ρ*). Equation 12 requires that the resulting risk function is normalized.

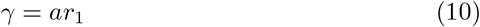

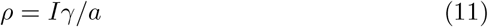

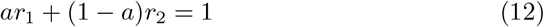

The parameters describing the piecewise function are then:

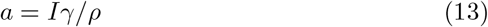

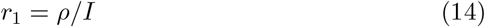

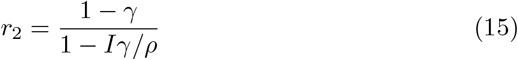

One additional requirement is imposed: if the filter is ‘worse’ than randomly guessing (precision *<* prevalence), then a uniform risk function is used instead (*r*_1_ = *r*_2_ = 1). In practice we likely would still want to use this information, but it would not provide a huge benefit, so this approximation is reasonable for an order-of-magnitude analysis. This requirement prevents nonsensical answers with *a >* 1 or *r*_1_ *< r*_2_.

With the risk function constructed, the minimum cost control policy can be evaluated for any combination of control, prevalence, sensitivity and precision. Equation 16 is the cost of the optimal solution when *Cost*(*d*) = *d* (Proof in Appendix D). This is the lowest achievable cost when reducing transmissions by fraction *β* using a filter with sensitivity *γ* and precision *ρ*.

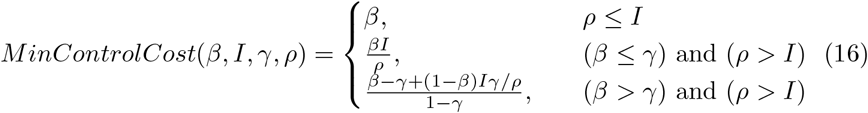

Figure 2 shows the minimum cost of control for filters with different precision and sensitivity. When the required level of control exceeds the sensitivity of the filter, the cost increases dramatically because the entire population must socially distance. The dependence on precision is much less sharp -so long as precision exceeds disease prevalence the filter provides some benefit. The bottom left region of each subplot in Figure 2 represents the second condition in Equation 16, in which both sensitivity and precision are adequate. In this region, 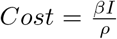, so the cost of the intervention is proportional to *I/ρ*. This means that the absolute value of precision is not important, but only the relative size compared to disease prevalence. So for example, a filter with a precision of 10^−5^ could be extremely useful when *I <* 10^−7^.

**Figure 2.**
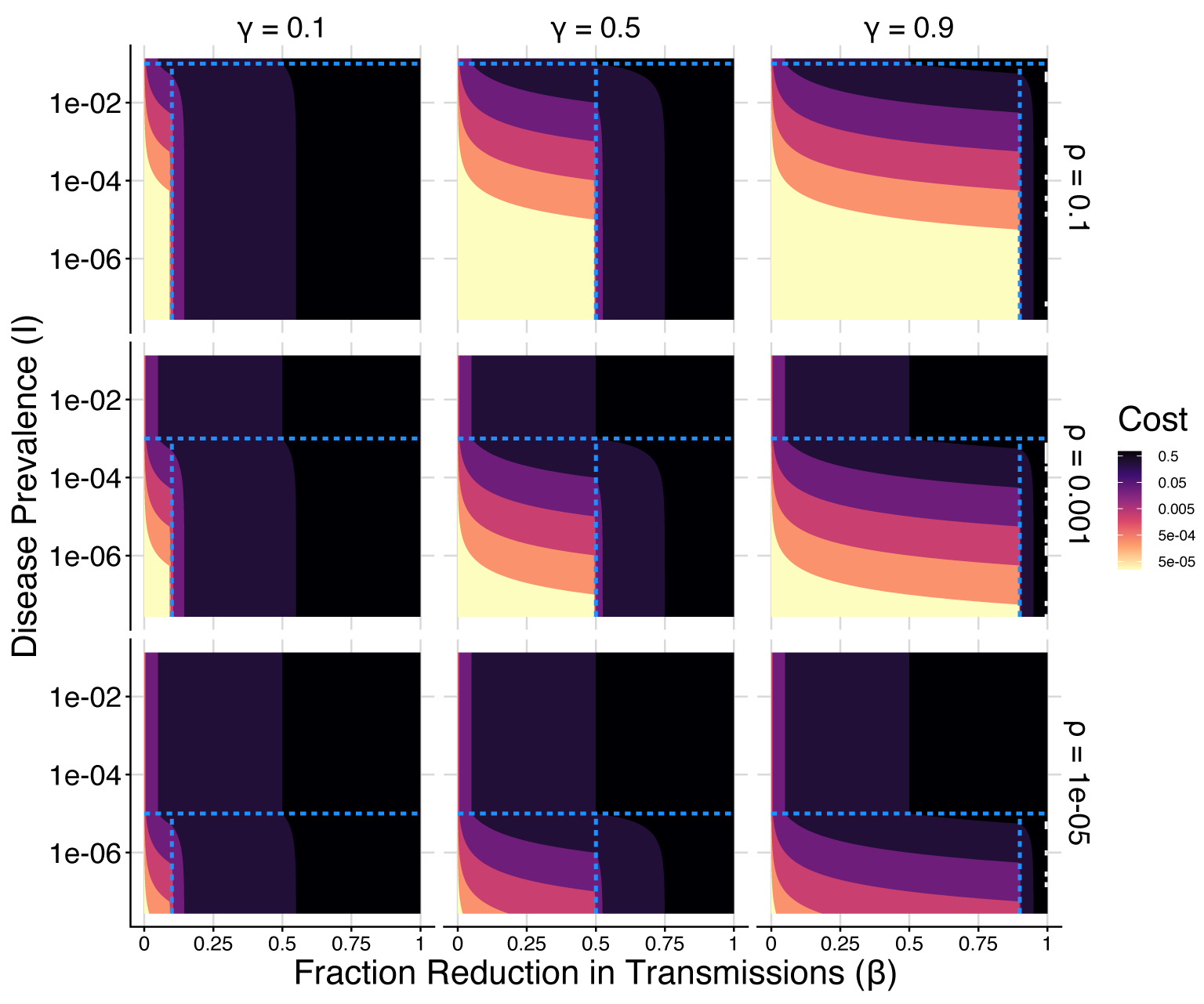
Minimum per-capita cost of control depending on required fractional reduction in expected transmissions (x axis) and disease prevalence (y axis). Results are shown for a linear cost function where total social isolation is a cost of 1. Horizontally, subpanels vary according to sensitivity *γ*, while vertically they vary according to precision . The horizontal line in each plot is set at the threshold where *ρ* = *I* (above which there is insufficient precising for targeting), and the vertical line is set so *β* = *γ* (to the right of which even perfect isolation of targeted individuals is insufficient without also introducing population-wide measures). Each of the sub-regions split by the dotted blue lines corresponds with a case in Equation 16.

## 6. Example Risk Functions: First and Second Degree Contact Tracing

We can similarly analyze the usefulness of empirically estimated infection risk among notified contacts using data from the NHS COVID-19 app published in Ferretti, Wymant et al. [12]. The data is split using a risk-scoring algorithm that divides each notified contact into one of 14 bins based on duration and proximity of exposure. The risk of infection is measured by recording the fraction of contacts within each bin that report a positive test after being notified^5^. This yields a piecewise constant (not-yet normalized) risk function, 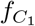(*q*) where *q* now represents quantile within contacts instead of within the population.

The information on risk among first degree contacts can be extrapolated to estimate infection risk amongst second degree contacts. This is useful to model whether it would be beneficial to expand to second degree contact tracing, an optional that the scaling offered by digital contact tracing makes much more practicable. The main motivation for considering second degree (forward) contact tracing is if the speed of transmission is so fast that by the time first degree contacts have been notified they may have already infected others ^6^. One solution is to make testing and contact tracing faster, but if that isn’t possible another option is to immediately notify second degree contacts.

Given the fraction of all infections that are detected as first 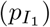 degree contacts, and that are detected as second degree 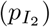 contacts, we can construct *f* (*q*) for the whole population. We assume that the risk distribution from detected first degree contacts is representative so that without interventions there are an average of *R*_0_*/E* [risk of infection | contact] contacts per infected individual. We assume for simplicity each contact is a unique individual. The approach used to create the combined risk function is described in Appendix E. Assuming that very reliable contact tracing is being done, but disease spread is quite fast (e.g. influenza with a generation time between 2-4 days [13]), we may prevent roughly half of transmissions via first degree tracing, so that second degree contacts would have the majority of the remaining infection risk. In this scenario (and assuming a very high test reporting rate), 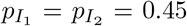 is a potential allocation of infection risk.

Figure 3 shows the risk quantile function using just first degree tracing 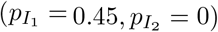 and both first and second degree tracing 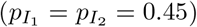. The highest risk individuals are first degree contacts, so the functions superimpose on the left. However, when low disease prevalence makes it useful to distinguish among lower risk individuals, second degree tracing makes this possible.

**Figure 3.**
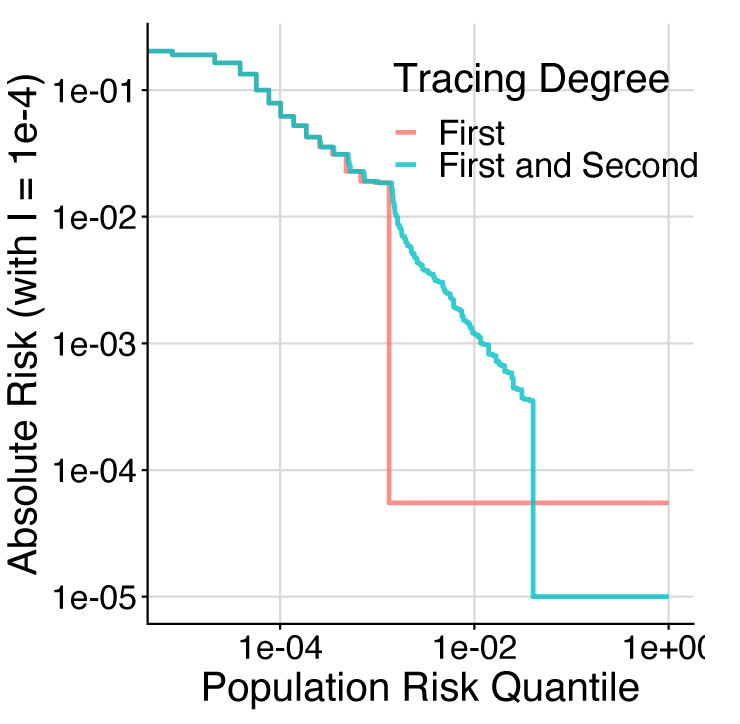
Risk function for first degree contact tracing (red) and both first and second degree contact tracing (blue). Due to assumption of non-clustered contact network structure, the addition of second degree tracing doesn’t change high-risk estimates. However, it provides significantly more information about moderate risk contacts. As discussed before, the value of this moderate-risk information depends on disease prevalence.

The minimum possible cost of achieving a fractional reduction in transmissions can be computed for these two risk quantile functions. With a linear social distancing cost function (*Cost*(*d*) = *d*), the minimum cost of control is the inverse of the integral of *f* (*q*) (as shown in Appendix A). Figure 4 is constructed using this approach, showing the minimum cost of control for different values of disease prevalence. Both sub-figures are similar when the required control is low, however the scenario with second degree tracing included is able to achieve higher control values at much lower cost when the disease prevalence (*I*) is less than 10^−4^.

**Figure 4.**
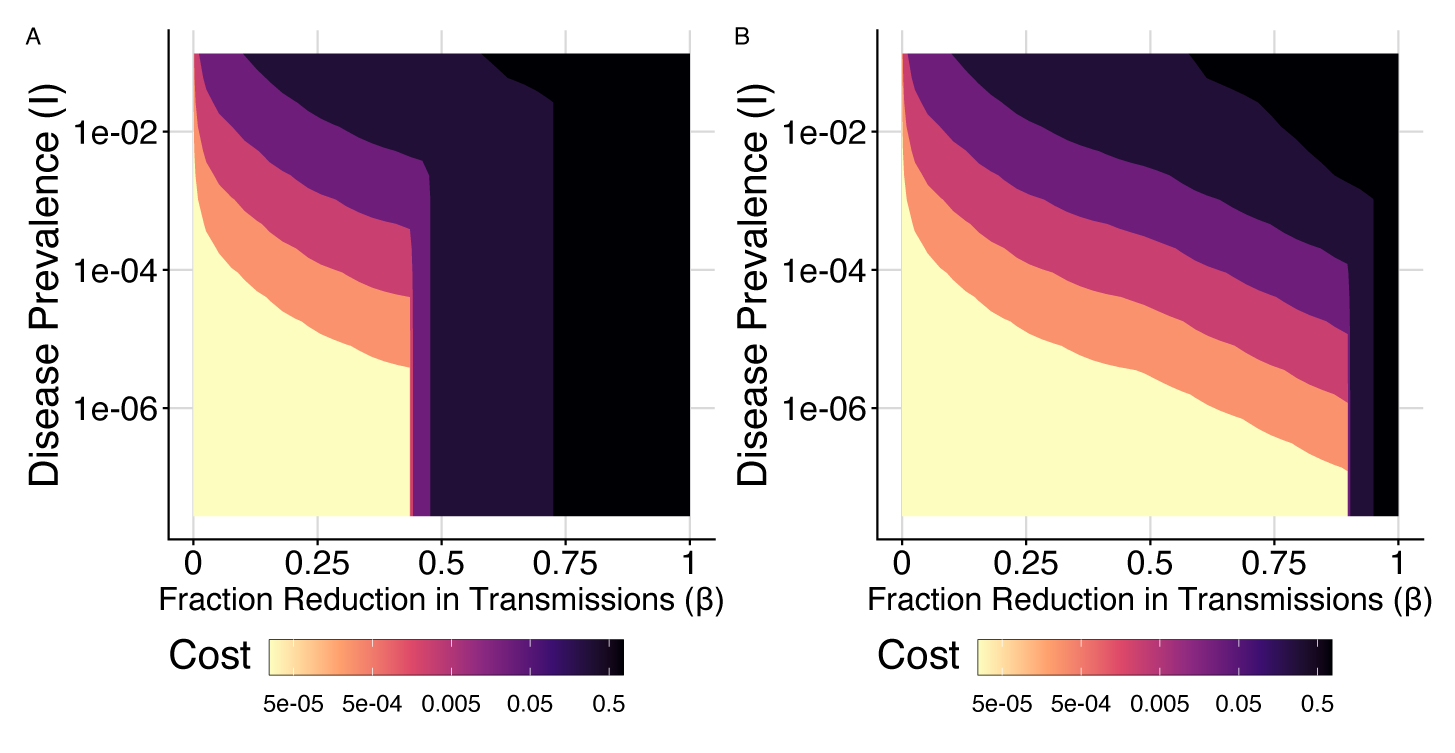
Minimum cost of control (for various values of fraction reduction in transmissions, *β*, and the fraction of the population infected, *I*) using the risk function from first degree contact tracing (A) and both first and second degree contact tracing (B). For low values of *β*, the cost is similar. For higher values of *β* and with *I <* 10^*−*3^, having access to risk information from second degree contact tracing dramatically lowers the cost of control.

## 7 Optimizing trade-offs between health and the burden of disease control

By assigning a cost to each infection, we can find the optimal control strategy that minimizes a weighted sum of control costs and infection costs by varying *β*. The cost per infection can be expressed in the same units as the cost of control using a metric like Quality Adjusted Life Years [9]. There may be uncertainty about the exact weights to assign to different outcomes, but these will likely be within an order of magnitude. The minimum possible cost of a disease trajectory (again assuming that infection-acquired immunity is negligible) from time 0 to T is then:

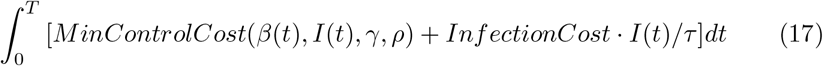

*I*(*t*) and *β*(*t*) are directly linked because the applied control influences the infection rate as shown in Equation 4.

### 7.1. Application: Steady Imported Cases

If possible, complete elimination of novel pandemics is often the most efficient option. However, complete disease elimination can be very difficult to achieve after substantial global spread. Strict border control to partially prevent local introductions of a pathogen is another option, although restricted travel does have a cost. The optimization problem with a variable importation rate is posed in Appendix F. For simplicity, in this section we will assume that the number of daily imported cases is fixed (this could happen if public health has limited control over movement into the region). Within this importation constraint, we can search for the best local disease control strategy.

We will further simplify the problem by restricting our search to (quasi) stationary solutions. We can find (quasi) stationary solutions by choosing *β* and *I* so the rate of change in Equation 4 is zero. With *A* and *I* set, the fractional reduction in transmissions (*β*) is constrained such that 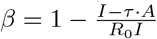.

The ongoing cost (per-person daily expected reduction in quality-adjusted-life from infection and disease mitigation) of a stationary solution can be calculated using Equation 17. Figure 5 shows an example of how cost depends on disease prevalence for a few scenarios with access to filters of different precision and sensitivity. In this example, *R*_0_ = 3, 100 cases are imported daily in a country with a population of 100 million, and the cost of each infection is 292 isolation days (equal to a 2% chance of a 20 year reduction in lifespan, then multiplied by 2 under the assumption that a day of full isolation is half the cost of dying a day earlier). At high disease prevalence, the expected cost of infection dominates the sum. As disease prevalence decreases, the cost of control remains similar but the cost of infection decreases, so the overall cost goes down. For each of the scenarios, when disease prevalence becomes lower than filter precision, the cost of control begins to decrease more rapidly. As the disease prevalence becomes quite low, the relative importance of the imported cases increases, so the fractional reduction in transmissions needs to be very high. The fractional reduction in transmissions needed for stability exceeds 1.0 when *I < A · τ*, which is not possible to achieve. For all of these curves, the ongoing cost can be minimized by choosing a disease prevalence that is set by the tradeoff between required control (which decreases with increasing prevalence), cost of control (which increases with prevalence at a rate depending on filter characteristics), and cost of disease (which obviously increases with prevalence).

**Figure 5.**
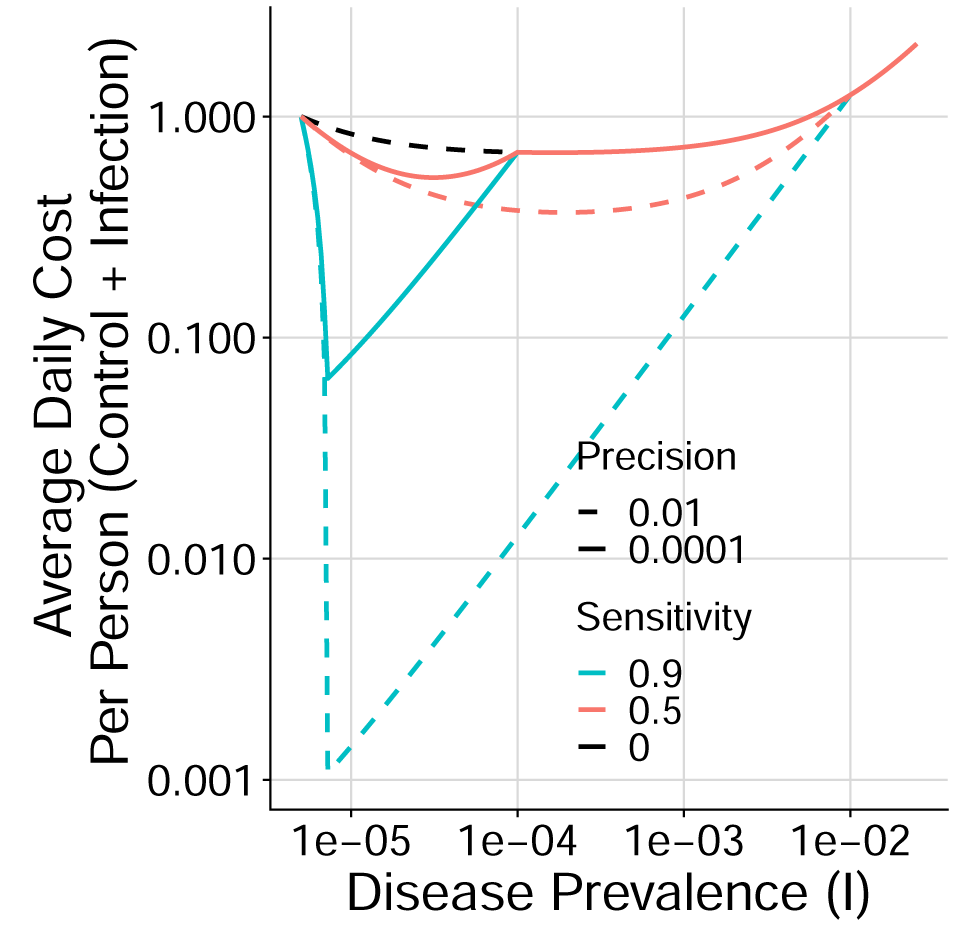
Daily per-person cost (in units of isolation-days) depending on disease prevalence, with control constrained so the system is stable (*dI/dt* = 0 in Equation 4) given imported cases. Each curve shows the minimum possible cost depending on the sensitivity and precision of the available filter. The curves differentiate when they reach the disease prevalence where the available precision becomes useful (precision greater than disease prevalence). High sensitivity filters (the two examples with 90%) are able to achieve low-cost equilibria near *I* = 1*e−*5. The cost of all scenarios increases at low disease prevalence because the relative influence of imported cases increases, meaning the fractional reduction in transmissions needs to be larger to maintain stability. At higher disease prevalence (*>* 1*/*1000), the daily cost increases because the cost of infection becomes significant.

This minimization problem is described by Equation 18. Figure 6 shows the minimum cost of stable disease control as precision and sensitivity vary for scenarios with different values of *R*_0_ and import rate (*A*). In most scenarios, if *γ >* (1 − 1*/R*_0_) and *ρ >* 200*Aτ*, then the per-person cost is less than 0.01 (in units of fraction of time in isolation). This is reasonable, because having a sensitivity greater than 1 1*/R*_0_ allows the disease to be controlled entirely with targeted interventions. Having a precision greater than 200*Aτ* means that with strong control, *I* could be held around 2*A · τ*, and the fraction of the population impacted by targeted interventions at a time would be less than 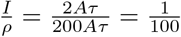

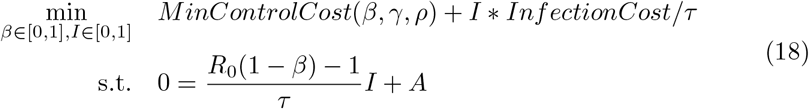

With a filter sensitivity large enough to independently control transmission with targeted interventions (*γ >* 1 − 1*/R*_0_), each introduction results in 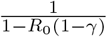additional infections, 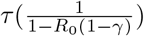 infectious days, and 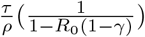 days of quarantine or isolation. With a reasonably good filter (e.g *ρ* = 1*/*100, *R*_0_(1 − *γ*) = 0.8), the cost per introduction could be on the order of 10^3^ or 10^4^ quarantine days. In a large country (e.g. with 10^8^ people), the per-capita cost per introduction could be on the order of 10^−4^ or 10^−5^ expected quarantine days. As an example, with *A* = 10^−6^ (100 introductions per day in a country of 100M), *τ* = 5, *R*_0_ = 3, *γ* = 0.8, and *ρ* = 1*/*100, the annual chance of infection is 1/1095, and the expected annual number of days in isolation is 0.45. In general, this means that while a large number of importations would require an unsustainable cost of control, with a moderate or low importation rate and a sufficiently strong filter, disease control would not significantly impact the average person.

**Figure 6.**
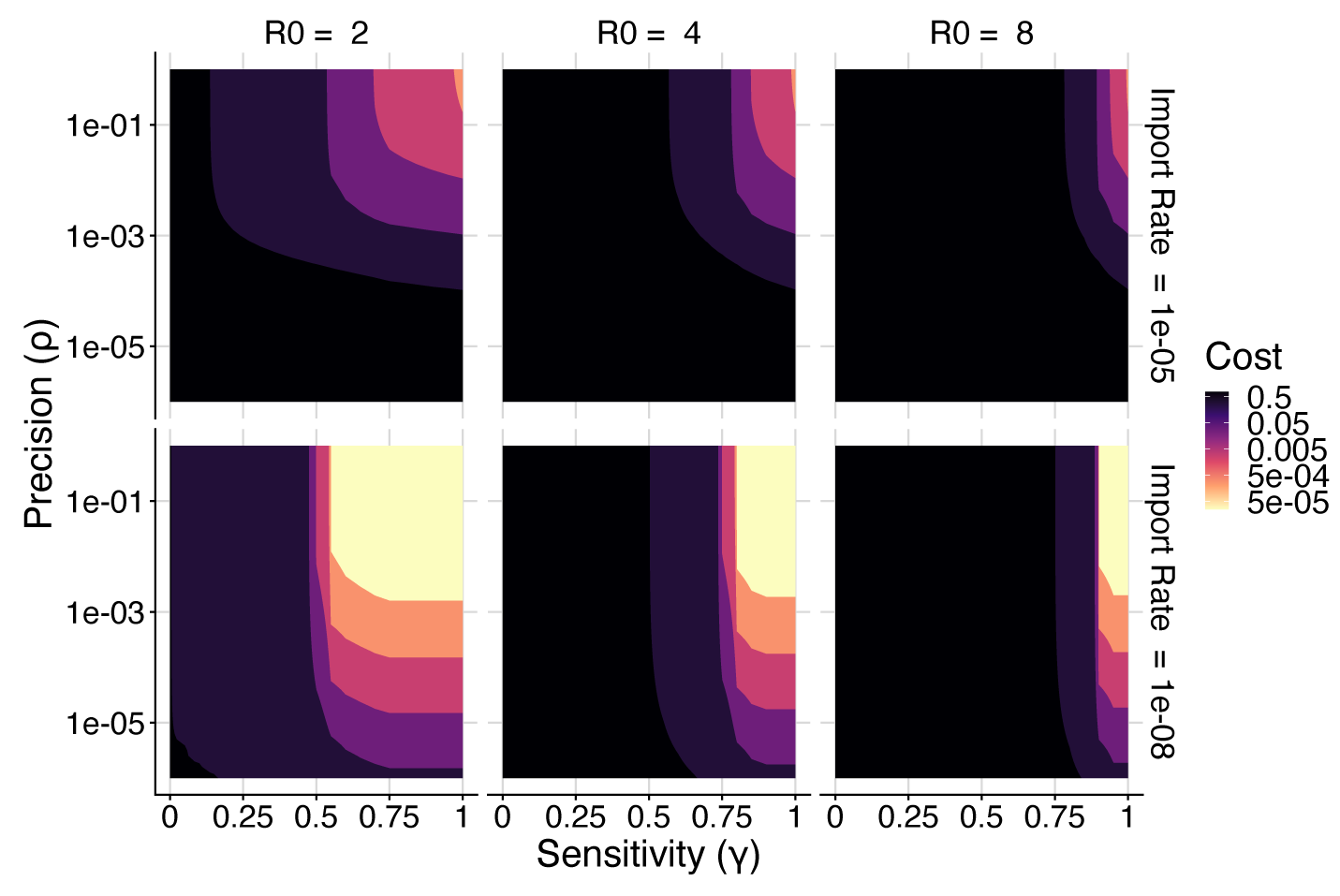
Minimum cost of stable (*dI/dt* = 0) control depending on filter precision and sensitivity. Each panel is for a scenario with different values of *R*_0_ and importation rate (*A*). The cost is in units of Quality Adjusted Days per person, and is the sum of the average burden imposed by disease control and risk adjusted cost of infection. The low cost region (Cost *<* 0.01) is roughly bounded by *γ >* 1 *−* 1*/R*_0_ and *ρ >* 200*Aτ* in each sub-figure.

## 8. Discussion

In this manuscript we developed computational tools to optimally target interventions based on risk of infectiousness. We showed that in some scenarios risk-targeted interventions can reduce the cost of disease control by several orders of magnitude compared to broad distancing. These methods can either be used to optimize interventions for specific infection risk and cost functions, or to provide more general recommendations based on a reasonable cost function and the summarized precision and sensitivity of a candidate filter.

In general, the cost of targeted interventions depends on precision relative to disease prevalence, not absolute precision^7^. Because of this, disease control gets much more efficient if the infection rate in the population can be reduced. The importation rate limits the minimum achievable disease prevalence because even with *R*_*t*_ below 1, imported cases can sustain the epidemic. If it is possible to control the importation rate, the optimal combined strategy can be computed as described in Appendix F. Even if the importation rate cannot be reduced to zero, low cost control can still be achieved with access to a sufficiently sensitive and precise filter.

High precision filters are preferable, but in some scenarios the ability to make use of lower precision information could enable disease control with targeted interventions where it was previously not possible. Lower precision information is often faster, more reliable, and cheaper to collect. For example, testing wastewater for every 10,000 homes is much easier to do than individually testing the entire population (especially in the beginning of a pandemic when testing capacity is lower).

Many existing strategies only roughly estimate infection risk. For order-of-magnitude estimates this is sufficient, but there is substantial room for refinement. Data on potential risk factors combined with test results can be used to train statistical models to much more accurately predict infection risk. As an example, Ferretti and Wymant et al [12] use data from the NHS app to train gradient-boosted trees to predict infection status based on duration and proximity of exposure. Isotonic regression [14] can be used to convert model prediction scores into calibrated probabilities.

The analysis in this manuscript is conditioned on the decision to perform highly successful disease control where the vast majority of the population is not infected. It would be prudent to separately compare the ongoing cost of control against alternatives like mass infection, including in the wake of the development of an only partially effective vaccine.

Even when only a small fraction of the infected population can be detected and convinced to isolate, targeting interventions based on infection risk can significantly reduce the cost of disease control by partially substituting for broad social distancing [8]. However, to reduce the cost by several orders of magnitude by eliminating the need for broad social distancing altogether, the sensitivity of targeted interventions must be greater than 1 − 1*/R*_0_. There are significant real world challenges associated with achieving this level of sensitivity, but some countries did demonstrate the success of targeting infection control based on low precision information during the COVID-19 pandemic [15]. Duplicating this approach in other countries is difficult because successful isolation of infectious individuals is crucial, but many are not comfortable strongly enforcing it. A potential solution is to strongly incentivize cooperation with important recommendations, which we hope to investigate in future work. Despite existing challenges, the potential for huge reductions in the cost of disease control motivates further investigation of risk-targeted strategies.

## Data Availability

All code and data used for this manuscript are available at https://github.com/JamesPetrie/RiskControl

## Acknowledgements

We would like to thank Luca Ferretti for valuable advice on risk analysis and the mathematical framing used in this manuscript.

## Appendices

## A Minimization with Linear Control Cost

For a linear (and positively sloped) control cost, *Cost*(*d*) = *A* ∗ *d* with *A >* 0, the problem in Equation 5 is equivalent to the problem in Equation 19.

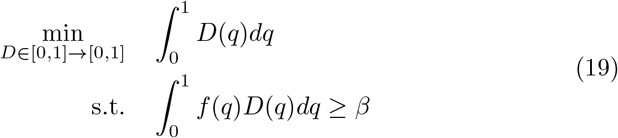

Here we will show that the optimal solution to Equation 19 can be found by solving the (easier) scalar optimization problem in Equation 20.

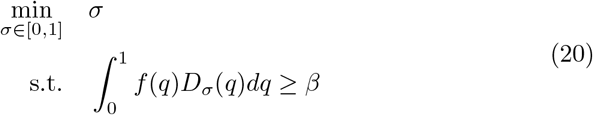

With:

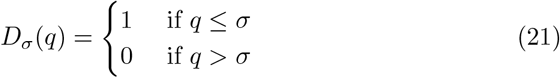

Proof ^8^:

First, we note that any solution, *D*_*σ*_(*q*), that is feasible for Equation 20 is also feasible for equation 19. Next we show that 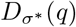 is optimal for Equation 20 then it is also optimal for Equation 19.

Equation 19 has convex constraints and a linear objective function, so it is a convex optimization problem. Therefore a locally optimal solution is also globally optimal (but not necessarily unique). We will show that the solution 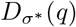 is locally optimal for Equation 19 by demonstrating that any feasible perturbations increase the objective value.

Let Ψ_*a,b*_ denote the set of functions with non-negative value, support [*a, b*], and that integrate to 1. Then all feasible perturbations of 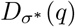 are contained in:

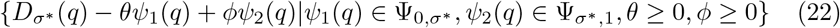

The optimal solution to 20, 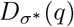, must have 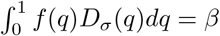 because if 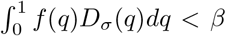 it would be infeasible and if 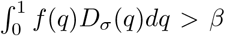 then the solution isn’t optimal because a lower objective value could be achieved by reducing *σ*. Since 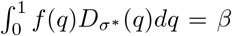, we must have 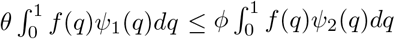 so that the perturbed solution still satisfies the constraint. Because the support of the two perturbation functions is known, the inequality can be rewritten as 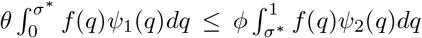. Since *f* (*q*) is monotonically decreasing:

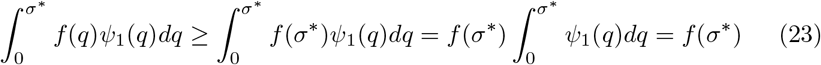

And

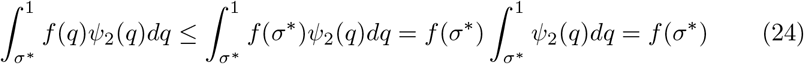

Therefore, for the perturbed solution to be feasible we require 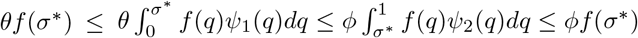 or more simply *θ* ≤ *ϕ*.

The change in objective value from the perturbation is 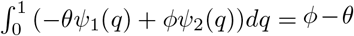. Because *θ* ≤ *ϕ* this means that for all feasible perturbations of 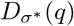 the objective function does not decrease. Therefore 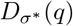 is locally and globally optimal for Equation 19.

### A.0.1 Rearranging results

Finally, we can further simplify the problem by rewriting Equations 20 and 21 as:

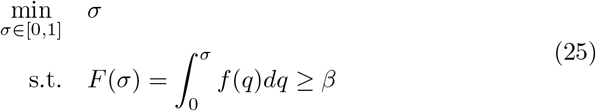

We’ve previously shown that the optimal solution to the problem must have *F* (*σ*^∗^) = *β*. While *F* (*q*) *<* 1 it must be strictly increasing because there is remaining risk so *F* ^′^(*q*) = *f* (*q*) *>* 0. Therefore, *F* (*q*) is invertible over the domain of interest and we can write *σ*^∗^ = *F* ^−1^(*β*). Therefore Equation 6 is equivalent to, 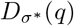.

## B Minimization with Strongly Convex Control Cost

When the function *Cost*(*d*) in Equation 5 is strongly convex, then the optimization over the space of functions *D*(*q*) can be replaced by the much easier optimization over a single scalar value *µ*_*β*_ in Equation 7.

Proof:

We will show Equation 7 is equivalent to Equation 5 for any discretization of *D*(*q*) and *f* (*q*) and take the limit as the discretization size goes to 0.

The discretized version of Equation 5 is:

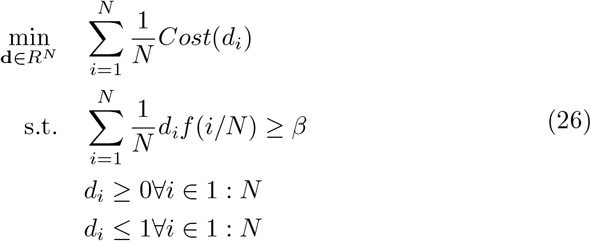

This is an optimization problem with a strongly convex objective and linear constraints defining a compact, non-empty feasible space. Therefore any vector satisfying the KKT conditions is globally optimal.

The KKT conditions for this problem are:

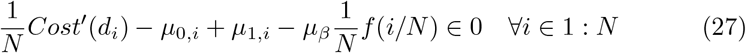

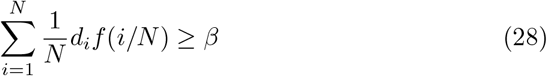

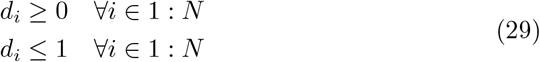

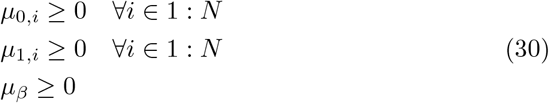

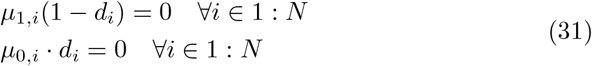

Here *Cost*^′^(*d*) is the one dimensional subdifferential of *Cost*(*d*) (which must exist because the function is strongly convex). *µ*_0_ corresponds to the dual variables for the ≥0 constraint, *µ*≤_1_ for the 1 constraint, and *µ*_*β*_ a single dual variable for the disease control constraint.

The simplification is possible because for any value of *µ*_*β*_ the vector **d** is uniquely defined. By leaving *β* unspecified for now and fixing *µ*_*β*_ we can compute this unique solution. Due to the complementary slackness constraints, *µ*_1,*i*_ is zero unless *d*_*i*_ = 1 and *µ*_0,*i*_ is zero unless *d*_*i*_ = 0. Each value of *d*_*i*_ will be either 0, 1, or a value between 0 and 1, so we can check each of these cases for equation 27 separately.

Case 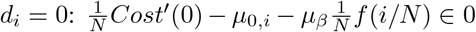 Because Cost(d) is a non-decreasing function^9^ *Cost*^′^(*d*) ≥ 0. *µ*_0,*i*_, *µ*_*β*_ and *f* (*i/N*) are all non-negative, so this equation is only be satisfied if 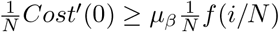).

Case 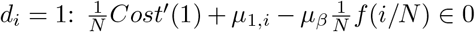. By similar reasoning, this equation is only satisfied if 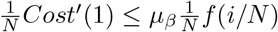

Case 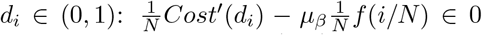. For this equation to be satisfied there must be some 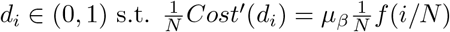.

Because *Cost*(*d*) is strongly convex, there are no two values *d*_*i*1_, *d*_*i*2_ with *d*_*i*1_ ≠ *d*_*i*2_ s.t. *Cost*^′^(*d*_*i*1_) = *Cost*^′^(*d*_*i*2_). Therefore for any *µ*_*β*_ and *f* (*i/N*), Equation 27 has the following unique solution.

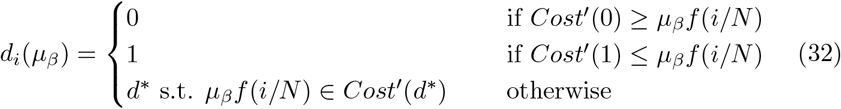

So for any value of *µ*_*β*_, there is a unique solution **d**(*µ*_*β*_). Each value *d*_*i*_ is monotonically increasing with *µ*_*β*_ so the fractional reduction in transmissions (*β*), and the total cost 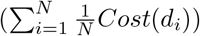 are also monotonically increasing with *µ*_*β*_. Because Equation 26 must have a unique solution, it is sufficient to find the smallest value of *µ*_*β*_ such that the solution vector **d**(*µ*_*β*_) satisfies the control constraint: 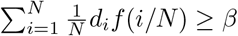. The simplified discretized problem is then:

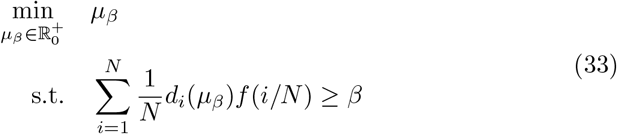

Taking the limit as *N* → ∞ yields Equation 7.

## C Minimization of Linear Control Cost with Piece-wise Constant Risk Function

For the case where the control cost is linear and the risk function is piecewise constant with two segments it is useful (and possible) to generate an explicit optimal solution.

In this case, *f* (*q*) is defined by Equation 9 (with 0 ≤ *a* ≤ 1 and 0 ≤ *r*_1_ ≤ *r*_2_ ≤ 1). Here, Equation 5 can be simplified to Equation 34.

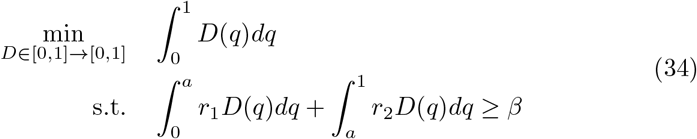

One optimal solution can be computed using the simplification from Equation 20. This yields the following solution:

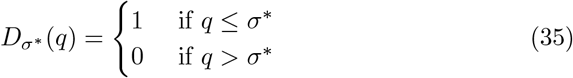

With:

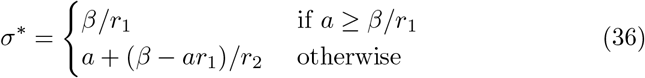

We will show there is another optimal solution for this case by constructing it and demonstrating that it is feasible and obtains the same objective value as solution 35.

This other optimal solution is:

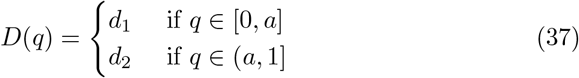

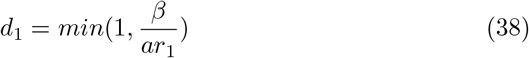

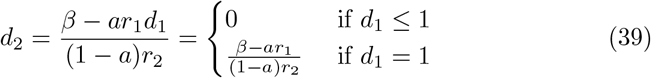

This solution essentially applies all control to the highest risk group until that saturates (at *d*_1_ = 1) and then applies the remaining control needed to the lower risk group.

Solution 37 satisfies the two constraints: *d*_1_, *d*_2_ ∈ [0, 1] if *β* ∈ [0, 1], and:

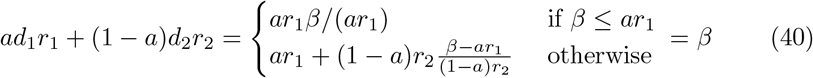

Solution 37 has an objective value of:

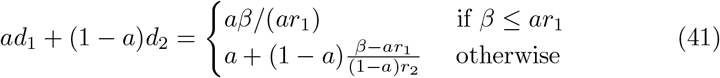

This objective value is identical to the one from solution 35 (which is just the value of *σ*^∗^). Because the proposed solution is feasible and attains the optimal objective, it is also optimal.

## D Simplified Optimal Solution Given Filter Precision and Sensitivity

The solution from the previous section can be used for the special case where the shape of the piece-wise risk function depends on filter sensitivity and precision as given by equations 13, 14, and 15 (and again using a linear cost function).

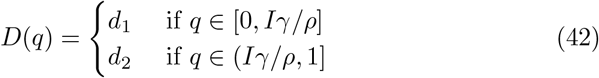

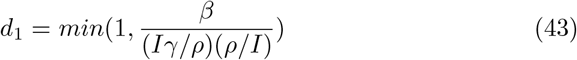

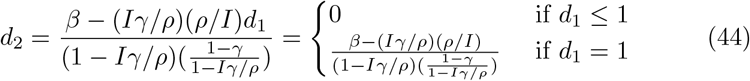

This solution can be simplified algebraically to yield the minimal objective value given by Equation 16.

## E Construction of whole-population risk quantile function with first and second degree tracing

The quantile function of second degree contact tracing risk 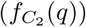 can be roughly constructed using 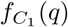. We assume that the contact network structure is tree-like, that is, there are no contacts that are notified of exposure multiple times. In reality, the contact network structure is likely to be much more clustered, with some people notified of exposure from multiple links; more sophisticated models attempt to account for this [16]. The risk function generated by the ‘tree-like’ assumption is the least-dense possible, and so it provides a lower bound on the usefulness of second degree contact tracing.

The risk of a second degree contact can be generated by taking the product of two independently generated first degree contact risks, as in Equation 45.

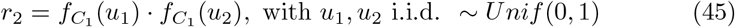

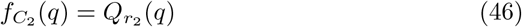

Then 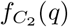 (*q*) is defined by Equation 46, where 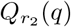 (*q*) is the quantile function applied to the random variable *r*_2_. This computation is fairly simple when dealing with binned risk data: the risk of a second degree contact is the product of two first degree risks, and the probability of that scenario is the product of each of the first degree probabilities. Ferretti, Wymant et al. [12] classified risk on the basis of duration of exposure and strength of Bluetooth signal recording between smartphones, and computed the probability of subsequently reporting a positive test for SARS-CoV-2. We discretized this into 14 first degree risk bins, yielding 196 second degree risk bins, which are then sorted in order of decreasing risk.

The constructed quantile functions are only for detected first and second degree contacts. To use the same approach for minimizing the cost of control from Section 4, the contact risk needs to be combined in one function, *f* (*q*), depending on the infection risk of the whole population. If we assume that 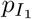 fraction of infectious individuals are identified as first degree contacts, 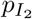 fraction are identified as second degree contacts, and the remaining 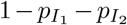 infection risk is evenly distributed amongst the whole population, then we can combine these quantile functions.

The fraction of the whole population that is identified as a first degree contact, 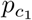, or second degree contact, 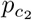, can be calculated using equations 47 and 48. These equations expand on the relation *P* (*C*_1_|*I*)*P* (*I*) = *P* (*I*|*C*_1_)*P* (*C*_1_).

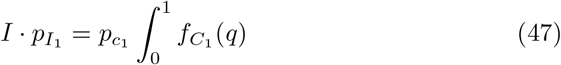

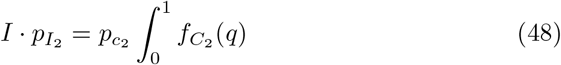

To normalize the risk functions, we divide by the disease prevalence *I*. Finally, we add in the remaining risk 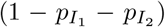 equally among the whole population, yielding equation 49.

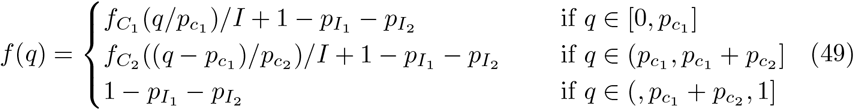

## F Further Optimizations: Border Policy and Dynamic Control

The optimization approach from the previous section can be extended to handle variable importation rates and dynamic disease control strategies. In this section we will introduce these problems at a high level.

Previously we assumed the importation rate of new cases was a fixed quantity. In reality, policy makers can influence this value through travel restrictions, testing requirements, quarantine, and other measures. If the cost of reducing the daily importation rate to a value *A* can be estimated, then we can represent this cost estimation with the function *BorderCost*(*A*). This function would likely have a very low cost at high values of *A*, the cost would increase dramatically as control measures begin to impact trade routes, and potentially diverge at low values due to very difficult to control undocumented border crossings. The overall (static) optimization problem is then given by Equation 50. Introducing more flexibility at worst leaves the minimum cost unchanged, but could potentially allow for much more efficient solutions by moving the importation rate to a level much lower than the precision of the available filter.

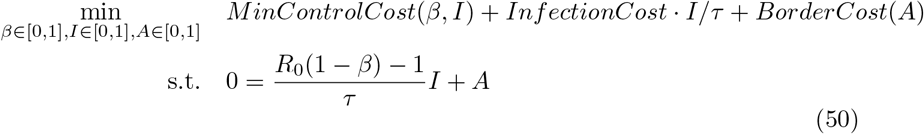

Finally, we can consider the optimization of dynamic disease trajectories. The (time-invariant) optimal solution can be found by solving for *β*(*I*) and *A*(*I*): functions that determine the disease control strategy depending on the current disease prevalence. This optimization problem is given by Equation 51.

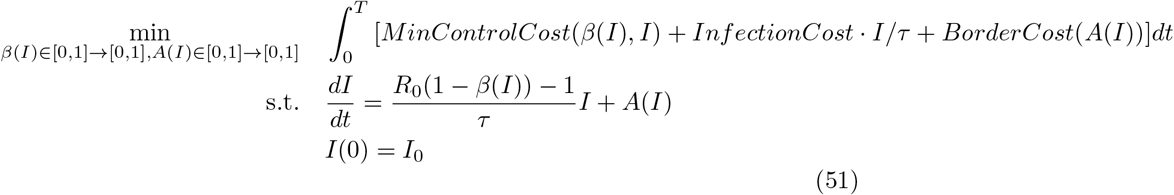

## Footnotes

1 1*−* 1*/R*_0_ = 0.66 with some overshoot

2 Calculated based on steady infection level of *I* = 5 *∗* (100*/*100*e*6 + 1*/*1000 *∗* 1*/*365) and social distancing set so derivative in Equation 4 is zero: 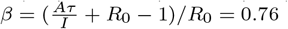

3 Valid if *I <* 1*/*1000 and *T <* 100*τ*, so that 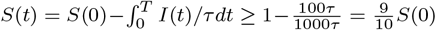

4 for the linear case the optimal strategy may not be unique

5 This is an underestimate of infection risk because not all infected contacts will be tested or test positive. Because this is an underestimate of infection risk it provides a lower bound on the usefulness of risk information from digital contact tracing

6 Backward contact tracing is not modeled here, but it can be used to alleviate difficulties with case detection

7 Similar to the dependence of the optimal risk threshold on disease prevalence in [8]

8 There is likely also a way to show this is as an application of Pontryagin’s maximum principle

9 it would not make sense for more distancing to reduce the cost

## Notes

### Competing Interest Statement

The authors have declared no competing interest.

### Funding Statement

This study did not receive any funding

## References

[1] Lin F, Muthuraman K, Lawley M. An optimal control theory approach to non-pharmaceutical interventions. BMC Infectious Diseases. 2010 2;10(1):1–13. Available from: https://link.springer.com/articles/10.1186/1471-2334-10-32https://link.springer.com/article/10.1186/1471-2334-10-32.

[2] Djidjou-Demasse R, Michalakis Y, Choisy M, Sofonea MT, Alizon S. Optimal COVID-19 epidemic control until vaccine deployment. Available from: 10.1101/2020.04.02.20049189.

[3] Perkins TA, España G. Optimal Control of the COVID-19 Pandemic with Non-pharmaceutical Interventions. Bulletin of Mathematical Biology. 2020 9;82(9):1–24. Available from: https://link.springer.com/article/10.1007/s11538-020-00795-y.

[4] Di Lauro F, Kiss IZ, Rus D, Della Santina C. Covid-19 and Flattening the Curve: A Feedback Control Perspective. IEEE Control Systems Letters. 2021 10;5(4):1435–40.

[5] Camelo S, Ciocan F, Iancu DA, Warnes XS, Zoumpoulis SI. Quantifying and Realizing the Benefits of Targeting for Pandemic Response. medRxiv. 2022 11:2021.03.23.21254155. Available from: https://www.medrxiv.org/content/10.1101/2021.03.23.21254155v5https://www.medrxiv.org/content/10.1101/2021.03.23.21254155v5.abstract.

[6] Birge JR, Candogan O, Feng Y. Controlling Epidemic Spread: Reducing Economic Losses with Targeted Closures. Management Science. 2022 3;68(5):3175–95. Available from: https://pubsonline.informs.org/doi/abs/10.1287/mnsc.2022.4318.

[7] Abdin AF, Fang YP, Caunhye A, Alem D, Barros A, Zio E. An optimization model for planning testing and control strategies to limit the spread of a pandemic – The case of COVID-19. European Journal of Operational Research. 2023 1;304(1):308–24.

[8] Petrie J, Masel J. The economic value of quarantine is higher at lower case prevalence, with quarantine justified at lower risk of infection. Journal of the Royal Society Interface. 2021;18(182):20210459.

[9] Torrance GW, Feeny D. Utilities and Quality-Adjusted Life Years. International Journal of Technology Assessment in Health Care. 1989;5(4):559–75. Available from: https://www.cambridge.org/core/journals/international-journal-of-technology-assessment-in-health-care/article/abs/utilities-and-qualityadjusted-life-years/AE0ECE718CD83E84569B54669D8D4006.

[10] Wallinga J, Lipsitch M. How generation intervals shape the relationship between growth rates and reproductive numbers. Proceedings of the Royal Society B: Biological Sciences. 2006 11;274(1609):599–604. Available from: https://royalsocietypublishing.org/doi/10.1098/rspb.2006.3754.

[11] Tian T, Huo X. Secondary attack rates of COVID-19 in diverse contact settings, a meta-analysis. The Journal of Infection in Developing Countries. 2020 12;14(12):1361–7. Available from: https://jidc.org/index.php/journal/article/view/13256.

[12] Ferretti L, Wymant C, Petrie J, Tsallis D, Kendall M, Ledda A, et al. Towards precision epidemiology: digital measurement of SARS-CoV-2 transmission risk. Manuscript submitted for publication. 2023.

[13] Böelle PY, Ansart S, Cori A, Valleron AJ. Transmission parameters of the A/H1N1 (2009) influenza virus pandemic: a review. Influenza and Other Respiratory Viruses. 2011 9;5(5):306–16. Available from: https://onlinelibrary.wiley.com/doi/full/10.1111/j.1750-2659.2011.00234.xhttps://onlinelibrary.wiley.com/doi/abs/10.1111/j.1750-2659.2011.00234.xhttps://onlinelibrary.wiley.com/doi/10.1111/j.1750-2659.2011.00234.x.

[14] Zadrozny B, Elkan C. Transforming classifier scores into accurate multiclass probability estimates. Proceedings of the ACM SIGKDD International Conference on Knowledge Discovery and Data Mining. 2002:694–9. Available from: 10.1145/775047.775151.

[15] Dao TL, Nguyen TD, Hoang VT. Controlling the COVID-19 pandemic: Useful lessons from Vietnam. Travel Medicine and Infectious Disease. 2020 9;37:101822. Available from: /pmc/articles/PMC7347475/ https://www.ncbi.nlm.nih.gov/pmc/articles/PMC7347475/.

[16] Guttal V, Krishna S, Siddharthan R. Risk assessment via layered mobile contact tracing for epidemiological intervention. medRxiv. 2020 5:2020.04.26.20080648. Available from: https://www.medrxiv.org/content/10.1101/2020.04.26.20080648v1https://www.medrxiv.org/content/10.1101/2020.04.26.20080648v1.abstract.

